# Ancestry Specific Polygenic Risk Score, Dietary Patterns, Physical Activity, and Cardiovascular Disease

**DOI:** 10.1101/2023.12.05.23299548

**Authors:** Dale S. Hardy, Jane T. Garvin, Tesfaye B. Mersha

## Abstract

**Background:** It is unknown whether the impact of high diet-quality and physical activity (PA) depends on the level of polygenic risk score (PRS) in different ancestries.

**Objective:** Determine the associations and interactions between high-risk PRSs, dietary patterns, and high PA with atherosclerotic cardiovascular disease (ASCVD) in European Americans (EAs) and African Americans (AAs). Another aim determined the molecular pathways of PRS-mapped genes and their relationships with dietary intake.

**Methods:** Cross-sectional analyses utilized de-identified data from 1987-2010 from 7-National Heart, Lung, and Blood Institute Candidate Gene Association Resource studies from the Database of Genotypes and Phenotypes studies for EAs (n=6,575) and AAs (n=1,606).

**Results:** The high-risk PRS increased ASCVD risk by 59% (Risk Ratio=1.59;95% Confidence Interval:1.16-2.17) in the highest tertile for AAs and by 15% (RR=1.15;1.13-1.30) and 18% (RR=1.18;1.04-1.35) in the second and highest tertiles compared to the lowest tertile in EAs. Within the highest PRS tertiles, high PA-diet combinations (Dietary Approaches to Stop High Blood Pressure (DASH), or Mediterranean, or Southern) reduced ASCVD risks by 9% (RR=0.91;0.85-0.96) to 15% (RR=0.85;0.80-0.90) in EAs; and by 13% (RR=0.87;0.78-0.97) and 18% (RR=0.82;0.72-0.95) for the DASH and Mediterranean diets, respectively in AAs. Top molecular pathways included fructose metabolism and catabolism linked to obesity, insulin resistance, and type 2 diabetes in both ancestries. Additional molecular pathways for AAs were Vitamin D linked to depression and aging acceleration; and death signaling associated with cancer.

**Conclusions:** Effects of high diet-quality and high PA can counterbalance the influences of genetically high-risk PRSs on ASCVD risk, especially in AAs.

## INTRODUCTION

Cardiovascular disease (CVD), a major cause of coronary heart disease (CHD) and stroke, is the leading cause of death in the US.^1^ Metabolic syndrome, an atherosclerotic disease precursor, defined as the clustering of ≥ 3 cardiovascular metabolic risk factors,^2^ more than doubles the risks of CHD, stroke, and cardiovascular mortality.^3^ Because atherosclerotic CVD (ASCVD) involves a prothrombic-and inflammatory state, efforts to decrease dyslipidemia and other metabolic derangements remains paramount. Several risk factors associated with ASCVD, and its complications include genetics, imbalanced diets, physical inactivity, and ancestry.^4^

Due to the lack of sufficient power to predict disease risk with clinical potential in individual patients from genome-wide association studies (GWAS), recent efforts have combined risk alleles and used their estimated allelic effects to create polygenic risk scores (PRS).^5^ However, because PRSs have been constructed primarily in populations of European ancestry to describe genetic risk, there is a need for studies that address the effects of PRS and diet on disease risks in minorities to increase generalizability in populations such as African Americans. In a PRS-type 2 diabetes study, a precursor disease to ASCVD, the highest risk for type 2 diabetes was seen in individuals with a high PRS burden and poor diet quality score.^6^ Irrespective of genetic risk, low diet quality was associated with approximately 30% increase in type 2 diabetes risk.^7^

Diets with high diet quality such as the Dietary Approaches to Stop High Blood Pressure (DASH) and Mediterranean diets, demonstrated evidence to reverse high blood pressure and dyslipidemia and reduce total mortality risk.^8,9^ The Mediterranean diet has been vastly recognized for its ability to decrease risks for CHD events, recurring CHD events, atherosclerosis CHD progression, sudden cardiac death, and all-cause mortality.^10–12^ The low-quality Southern diet has been associated with higher risk for CHD events^11^ and all-cause mortality.^13^ African Americans have fewer ideal health behaviors that promote cardiovascular health than European Americans, including lower intake of fruits and vegetables but higher red and processed meat intake, and higher rates of physical inactivity.^14,15^ Moderate to high physical activity, performed weekly with 150 to 300 minutes of moderate-intensity, or 75 to 150 minutes of vigorous-intensity aerobic physical activity, can reduce the risk of developing ASCVD, metabolic syndrome and other cardiometabolic conditions.^16^ Because ASCVD is a highly polygenic disorder that may differ by ancestry, understanding the molecular mechanisms linking genes with related ASCVD SNPs may give more insight into the pathways involved. Our main study aim was to investigate the associations and interactions between a genetically high-risk PRS, diet patterns, and high physical activity level with ASCVD risk by ancestry. A secondary aim examined the molecular pathways associated with the interrelated PRS-mapped genes.

## METHODS

Data available upon request from database of Genotypes and Phenotypes (dbGaP).^17^

### Selection of participants

Participants were obtained from seven parent studies from the National Heart, Lung, and Blood Institute (NHLBI) Candidate Gene Association Resource (Care) that are part of dbGaP data collection. We merged data from the Atherosclerosis Risk in Communities (ARIC) study,^18^ Coronary Artery Risk Development in Young Adults (CARDIA) study,^19^ Cardiovascular Heart Study (CHS),^20^ Framingham Heart Study (FHS) Offspring and GENX 3 studies,^21^ Multi-Ethnic Study of Atherosclerosis (MESA) study,^22^and Women’s Health Initiative (WHI) study,^23^ to create a large dataset by ancestry. All studies are large-scale, ongoing prospective cohort studies with atherosclerosis outcomes. Further design and sampling are explained elsewhere for all studies.^18–23^ Data collection years spanned from 1987 to 2010. All participants signed an informed consent prior to participation in the parent study. Morehouse School of Medicine Social & Behavioral Institutional Review Board approved the current study.

### Constructing the PRS and principal components for stratification

We performed SNP imputation using the Michigan Imputation Server algorithm using 1000 Genomes Phase 3 (Version 5).^24^ After quality cleaning and merging of imputed datasets, SNPs for the PRS were extracted using Plink, a whole genome association analysis toolset by ancestry.^25^ For SNPs within high linkage disequilibrium ≥ .8, tag SNPs were chosen based on higher binding capacity in RegulomeDB.^26^ The Hardy-Weinberg test for all SNPs was performed in Plink^25^ using chi-square goodness-of-fit test for European Americans and African Americans separately. The 10 genetic principal components were computed in Linux using gcta64 guidelines^27^ in RStudio^28^ to calculate a genetic-related matrix by ancestry and then specifying the principal components.

We chose metabolic syndrome SNPs with p < .05 for ASCVD by ancestry (European American or African American). We flipped alleles for SNPs that had reduced ASCVD risks making their effects risk-raising. We used a lower RegulomeDB score that was more predictive at least ≤ 4, which represented any transcription factor binding, matched motif, DNase peak up to any SNPs with only transcription factor binding plus DNase peak, but no motif.^26^ RegulomeDB presents a scoring system with functional categories ranging from 1 to 6 by the way of integrated annotations data on methylation, chromatin structure, protein motifs and binding. The lower the RegulomeDB score, the stronger the evidence for a variant to be in a functional genomic region. We randomly divided the combined dataset into train and test sets by ancestry in a 50/50 ratio. The PRS was trained using a metabolic syndrome-ASCVD phenotype. Construction of the ancestry specific PRSs were calculated by computing the sum of risk alleles corresponding to dietary patterns and cardiovascular disease, weighted by the effect size estimate of the risk variants identified from our genome-wide association studies. To increase the predictive power of the PRS, the joint power of multiple SNPs was included in the PRS using the summary best linear unbiased prediction (SBLUP) method. Then we generated summary statistics on the train dataset and replicated these statistics in the test dataset to create our high-risk PRS to use in our statistical models.

### Study Variables

We defined ASCVD, the outcome, as present if patients had a diagnosis of CHD, heart attack, stroke, trans-ischemic attack (TIA), or peripheral vascular disease. We used the first visit available as the baseline that had nutrition and physical activity data. ASCVD and all covariates were measured at the study baseline for: ARIC, CARDIA, CHS, and WHI were taken from Visit 1; for FHS Offspring and FHS GENX 3, data were taken from Exam 3 and Exam 2 respectively; and for MESA, data were taken from Exam 5. Covariates considered for adjustment were age (dichotomized), sex, physical activity (high/low), current smoking (yes/no), current drinking (yes/no), and total caloric intake.

### Data harmonization

We harmonized our data by bringing together data of varying formats (file formats, variable definitions, etc.) from the seven NHLBI Care datasets to generate a large cohesive dataset. We assessed all chosen variables to ensure their presence in all NHLBI Care datasets. Some variables were transformed to yes/no status to harmonize measures across datasets. For example, the physical activity variable had different formats across datasets. To harmonize the physical activity variable across datasets, we recoded this variable in high/low form by evaluating its functional form in each dataset. When we selected study variables that were present in all 7-NHLBI Care datasets, we created variable definitions by using existing variable definitions that agree across all datasets, so our chosen variables had consistent meaning across all datasets.

### Food frequency questionnaire assessment from multiple studies

Studies that used a semi-quantitative food frequency questionnaire (FFQ) to obtain information on dietary intake were ARIC,^29^ MESA,^30^ Framingham Heart Studies,^31^ and Women’s Health Initiative Study.^32^ Studies that used diet history recalls were CARDIA^33^ and CHS.^34^ Supplemental Table 1 shows more details of these studies. Generally, in the FFQ, participants reported their intake based on 9 levels of frequency, ranging from < 1 time per month to ≥ 6 times per day. In the diet history sessions, participants were asked questions about usual intake. At the examination, interviewers showed participants standard serving sizes, typical servings using food models, and additional information such as brand names of prepared foods to help them estimate intake.

### Dietary patterns construction from multiple studies

Dietary patterns were created for the DASH, Mediterranean, and Southern diets. These diets were constructed within each dataset from energy-adjusted foods and nutrients available from the FFQ or diet history. The DASH diet was constructed with nutrients such as Vitamin D, calcium, magnesium, potassium, phosphorus, potassium to sodium ratio, thiamine, niacin, and fiber that are integral parts of the benefits in lowering blood pressure.^35^ The Mediterranean diet was created with whole foods that have been proven to lower ASCVD risk, such as, fruits, vegetables, salads, nuts, wholegrains, beans and peas, dietary fiber, lean white meats, and fatty fish.^10,11^ The Southern diet was constructed with foods typical of a Western diet such as fried foods (French fries, fried chicken), sugar sweetened beverages (sodas, tang), chips, red meats (hamburgers, pork), processed meats (deli meats, ham, hotdog, salami), organ meats (liver, kidneys), eggs, French fries, alcohol, sweets and desserts.^10–13^ The healthy DASH and Mediterranean diets represented diets with high-quality diet scores, and the harmful Southern diet, a low-quality diet score. After we assembled our list of foods for each diet score, we grouped our observations using KMeans Cluster analysis.^36^ The KMeans Clustering algorithm finds observations that are like each other and places them into groups. All groups have minimum within-cluster variance, so observations within each group have similar characteristics.

### Statistical Analysis

We imputed missing observations on cigarette smoking, drinking status, and physical activity to increase our sample size especially for African Americans. Imputations were <5% of the original participant sample. Our initial sample without duplicates was based on the PRS (n=11,266). We excluded participants if they had missing observations at baseline on ASCVD and the covariate propensity score (n=2,982), total caloric intake < 600 Kcal or > 4,200 Kcal per day for men (n=55), and < 500 Kcal or > 3,600 Kcal per day for women (n=48). Our final models included 8,181 participants in which 6,575 (80.37%) were European Americans and 1,606 (19.63%) were African Americans. We did not drop missing observations across dietary patterns, because observations in dietary patterns were missing at random across dietary patterns and would have decreased the sample size further within each dietary pattern.

We created a principal component adjusted PRS. In models that evaluated the effects of physical activity and the PRS and dietary patterns on ASCVD risk, the physical activity variable was not included in the covariate propensity score. To conserve on power (especially for African Americans) and to decrease bias, we computed a covariate propensity summary score by ancestry by regressing ASCVD on the covariates. Generalized linear models (GLM) were used to derive risks ratios and 95% confidence intervals. In our GLM analysis, we were interested in the expectation of the outcome, ASCVD, as a function of the PRS and/or specific dietary pattern (DASH, Mediterranean, Southern) adjusted for the covariate propensity score.

In our multivariable models, we regressed ASCVD on the PRS and each dietary pattern, adjusting for the covariate propensity score. In interaction models, we included the PRS, each dietary pattern, along with the covariate propensity score for adjustment. All analyses were performed using tertiles of each dietary pattern by PRS tertiles. In all models, a 2-sided p < .05 and more stringent, Bonferroni adjustment for multiple testing (p <.025) were used as the threshold for statistical significance. All regression analyses were bootstrapped 10,000 times. Our multivariable statistical analyses were conducted using Stata MP, version 17.0 (StataCorp, College Station, TX).^37^

### Interaction between the PRS, dietary patterns, and physical activity on ASCVD risk

In interaction analysis, we were interested in additive interaction in which the modifying effect of the dietary patterns elicit changes on different levels of the PRS, and how high physical activity affected this relationship. We computed the difference in probability for ASCVD between the expected risks at lowest, second and highest tertiles of the PRS with each dietary pattern in the presence of high and low physical activity adjusting for the covariate propensity score.

### Pathway analysis of SNPs in the PRS mapped to genes

We mapped the SNPs to their respective genes by ancestry, using snpXplorer.^38^ SnpXplorer is a web-based application to explore human SNP-associations and annotate SNP-sets. Then we then took our gene list by ancestry and analyzed the genes by pathways in the Enrichr program.^39^ Enrichr uses a set of Entrez gene symbols as input. We presented visualizations using the bar graphs. The length of the bar represents the significance of gene-set. The brighter the color, the more significant the p value for the term.

## RESULTS

### Descriptive characteristics by ancestry

In this cross-sectional study, there were 8,181 participants: 6,575 (80.37%) European Americans and 1,606 (19.63%) African Americans. Table 1 shows the descriptive characteristics of the sample by ancestry. Unlike European Americans, a higher percentage of African Americans were more obese, were less physically active, had higher waist circumference, had higher systolic blood pressure, a higher proportion were on blood pressure medications, had lower HDL cholesterol, and had higher blood glucose levels. However, a higher percentage of European Americans drank more alcohol, had higher triglyceride levels, and more ASCVD events as explained by an increase rate of CHD events, stroke/TIA, and peripheral vascular disease.

**Table 1.** Characteristics among European Americans and African Americans at baseline.

|  | Ancestries<br>Combined | European<br>Americans | African<br>Americans | P Value Comparing<br>African Americans to<br>European Americans |
| --- | --- | --- | --- | --- |
|  | (n=8,181) | (n=6,575) | (n=1,606) |  |
| Characteristic | Mean (SD) or Column Percent (%) of Participants |  |  |  |
| Age (range:19-98 years; mean: 60 years; SD:10.5) |  |  |  | < .0001 |
| 20-57 years | 46.6 | 44.1 | 57.0 | <.0001 |
| 58-94 years | 53.4 | 55.9 | 43.0 |  |
| Female (%) | 50.8 | 49.5 | 56.3 | < .0001 |
| Body mass index (BMI) | 27.6 (5.1) | 26.9 (4.7) | 29.6 (6.0) | < .0001 |
| Weight status (%) |  |  |  | < .0001 |
| Underweight (BMI ≤ 18.5 kg/m<sup>2</sup>) | 1.0 | 1.0 | .9 |  |
| Normal weight (BMI 18.5-24.9 kg/m<sup>2</sup>) | 33.0 | 36.4 | 17.1 |  |
| Overweight (BMI 25.0-29.9 kg/m<sup>2</sup>) | 41.7 | 41.3 | 43.4 |  |
| Obese (BMI ≥ 30 kg/m<sup>2</sup>) | 24.7 | 21.3 | 38.7 |  |
| High physical activity level (%) | 51.3 | 51.9 | 48.4 | .012 |
| Current cigarette smoking (%) | 32.7 | 32.6 | 32.9 | .828 |
| Current alcohol intake (%) | 53.3 | 57.3 | 37.2 | < .0001 |
| Waist circumference (% > 102 cm: men and > 88cm: women) | 65.7 | 64.4 | 71.0 | < .0001 |
| Systolic blood pressure (% > 120mmHg) | 55.1 | 53.1 | 63.4 | < .0001 |
| HDL (% < 40mg/dL: men and < 50mg/dL: women) | 65.3 | 65.2 | 65.7 | .703 |
| Triglycerides (% > 150mg/dL) | 28.1 | 31.0 | 16.4 | <.0001 |
| LDL (% > 100 mg/dL) | 80.7 | 81.0 | 79.5 | .158 |
| Fasting blood glucose (% > 100 mg/dL) | 48.6 | 48.2 | 50.6 | .076 |
| Blood pressure medications (%) | 36.5 | 33.4 | 49.1 | < .0001 |
| Cardiovascular disease (%) | 17.0 | 18.0 | 13.0 | < .0001 |
| Coronary heart disease (%) | 10.4 | 11.6 | 5.6 | < .0001 |
| Stroke/TIA (%) | 8.1 | 8.6 | 6.1 | < .0001 |
| Peripheral vascular disease (%) | 2.9 | 2.6 | 4.4 | .001 |

**Abbreviations:** HDL, high density lipoprotein cholesterol; LDL, low density cholesterol; TIA, trans ischemic attack; sd, standard deviation %, percent of sample. All other variable results are means.
P values for proportions of categorical variables comparing African Americans to European Americans were calculated using Pearson's chi-square tests of hypothesis for independence or t-tests for continuous variables to compare equality between means between ancestries. Analysis showed that all variables were statistically significant between each other except current cigarette smoking, HDL cholesterol, LDL cholesterol, and fasting blood glucose.

### The PRS was associated with ASCVD risk by ancestry

The genetically high-risk PRS were associated with ASCVD. African Americans had higher magnitude of associations than European Americans (Table 2). In the continuous PRS, each unit in Z-score per 1-SD increase was associated with an 8% higher ASCVD risk in European Americans (Risk Ratio (RR)=1.08; 95% Confidence Interval:1.03-1.13) and a 23% higher risk (RR=1.23;1.08-1.40) in African Americans. PRS in higher tertiles vs. the lowest tertile differed in ASCVD risks. Among European Americans, we found a 15% higher risk for ASCVD (RR=1.15;1.13-1.30) in tertile 2 vs. lowest tertile; and 18% higher ASCVD risk (RR=1.18;1.04-1.35) in the highest tertile compared to the lowest tertile. For African Americans, we observed 59% higher ASCVD risk (RR=1.59;1.16-2.17) for highest tertile compared to lowest tertile. All effect estimates mentioned, passed Bonferroni cut-off for false discovery rate at p < .025.

**Table 2.** Association between a polygenic risk score, dietary patterns, physical activity, and cardiovascular disease at baseline.

|  | Risk Ratio (95% Confidence Interval) P Value |  |  |  |
| --- | --- | --- | --- | --- |
|  | European Americans<br>(n=6,575) | P Value | African Americans<br>(n=1,606) | P Value |
| **Polygenic risk score** |  |  |  |  |
| **PRS per 1 SD Z score** | **1.08 (1.03-1.13)** | **.003\*** | **1.23 (1.08-1.40)** | **.001\*** |
| PRS lowest tertile | 1.00 (ref) |  | 1.00 (ref) |  |
| PRS second tertile | **1.15 (1.13-1.30)** | **.031** | 1.33 (0.96-1.46) | .085 |
| PRS highest tertile | **1.18 (1.04-1.35)** | **.009\*** | **1.59 (1.16-2.17)** | **.005\*** |
| **Dietary Patterns** |  |  |  |  |
| **DASH diet per 1 SD Z score** | **0.92 (0.88-0.97)** | **.002\*** | **0.83 (0.73-0.94)** | **.004\*** |
| Lowest tertile | 1.00 (ref) |  | 1.00 (ref) |  |
| Second tertile | 0.94 (0.83-1.06) | .320 | 0.80 (0.60-1.07) | .141 |
| Highest tertile | **0.85 (0.75-0.95)** | **.006\*** | **0.64 (0.47-0.89)** | **.008\*** |
| **Mediterranean diet per 1 SD Z score** | **0.90 (0.85-0.95)** | **<.0001\*** | **0.87 (0.77-0.99)** | **0.033** |
| Lowest tertile | 1.00 (ref) |  | 1.00 (ref) |  |
| second tertile | **0.85 (0.76-0.96)** | **.011\*** | 1.13 (0.65-1.97) | .668 |
| highest tertile | **0.83 (0.72-0.95)** | **.008\*** | **0.71 (0.52-0.97)** | **.029** |
| **Southern diet per 1 SD Z score** | **1.08 (1.02-1.14)** | **.004\*** | **1.25 (1.08-1.44)** | **.002\*** |
| Lowest tertile | 1.00 (ref) |  | 1.00 (ref) |  |
| second tertile | **1.18 (1.04-1.33)** | **.009\*** | 1.31 (0.91-1.87) | .142 |
| highest tertile | **1.19 (1.05-1.34)** | **.008\*** | **1.76 (1.21-2.56)** | **.003\*** |
| **Physical activity** |  |  |  |  |
| Physical activity (high vs. low) | **0.71 (0.63-0.79)** | **<.0001\*** | **0.53 (0.40-0.69)** | **< .0001\*** |

**Bold indicates p values that were statistically significant at $p < .05$ .**
**\*Bonferroni adjustment for multiple testing ( $p < .05/2 = < .025$ ).**
PRS: Atherosclerosis cardiovascular disease status was regressed against the principal components adjusted PRS adjusting for a covariate propensity score composed of age, sex, physical activity, current cigarette smoking status, current drinking status.
Physical activity: Atherosclerosis cardiovascular disease status was regressed against physical activity adjusting for a covariate propensity score composed of age, sex, current cigarette smoking status, and current drinking status.

### Dietary Patterns were associated with ASCVD by ancestry

Table 2 shows the association of different dietary patterns on ASCVD by ancestry after adjusting for all covariates (age, sex, physical activity, current drinking status, current smoking status, and total caloric intake). Across all dietary patterns, African Americans had lower risks with the DASH and Mediterranean diets, and higher risks with the Southern diet than European Americans. For each unit increase in Z-score, the DASH score showed 8% lower risk for European Americans but had 17% lower risk for African Americans. When we compared DASH diet tertiles, only the highest tertile compared to the lowest tertile were significant and met Bonferroni correction (p < .025). In European Americans, the DASH score showed 15% lower ASCVD risk (RR=0.85;0.75-0.95), and for African Americans, a 36% lower ASCVD risk (RR=0.64;0.47-0.89). African Americans had lower ASCVD risks with the Mediterranean diet than European Americans, but these results did not meet Bonferroni correction for multiple testing (p < .025). European Americans had 10% lower ASCVD risk per each unit higher in Z-score with the Mediterranean diet. European Americans had 15% lower ASCVD risk for second tertile compared to lowest tertile (RR=0.85;0.76-0.96) and 17% lower risk for highest tertile compared to lowest tertile (RR=0.83;0.72-0.95) in the Mediterranean diet. The Southern diet showed higher ASCVD risk in both ancestry groups. However, African Americans showed a higher magnitude of risk per 1 SD Z-score increase (RR=1.25;1.08-1.44) compared to European Americans (RR=1.08;1.02-1.14); While African Americans had a 75% higher ASCVD risk in the highest tertile compared to the lowest tertile (RR=1.76;1.21-2.56). European Americans had significantly higher risks but to a lesser magnitude in the second and (RR=1.18;1.04-1.33) and highest tertiles compared to the lowest tertile (RR=1.19;1.05-1.34).

### Physical activity was associated with ASCVD risk by ancestry

Table 2 shows the effects of physical activity on ASCVD risk after adjusting for covariates. Again, African Americans benefited from physical activity more than European Americans as indicated by a lowering of 47% in ASCVD risk (RR= 0.53;0.40-0.69) compared to European Americans who had a lowering of 29% in ASCVD risk (RR=0.71;0.63-0.79) after adjusting for covariates. ASCVD risks by ancestry met Bonferroni correction for multiple testing (p < .025).

### Dietary patterns and physical activity combined effects on ASCVD risk by ancestry

Supplemental Figure 1 shows the effects of each dietary pattern in the presence of physical activity on ASCVD risk by ancestry. In these models, we used the expected risk of consuming a specific diet with higher tertiles compared to the lowest tertile. In European Americans, we found highly significant effects (p < .0001) in the second and highest tertiles compared to the lowest tertile. The DASH, Mediterranean diets, and even the Southern had lower ASCVD risks with high physical activity level among European Americans in the range of 10% to 13%. However, among African Americans, we observed significantly lower ASCVD risks in the range of 6% to 10% for the combination of high physical activity with the DASH (RR=0.94;0.89-0.99) and Mediterranean (RR=0.90;0.86-0.95) diets, only in the highest tertile compared to the lowest tertile. Furthermore, among African Americans, we observed a 7% lower ASCVD risk with high physical activity’s influence over the low-quality Southern diet in the second tertile compared to the lowest tertile (RR=0.93;0.88-0.99). These results met Bonferroni adjustment for multiple testing (p <.025).

### PRS and physical activity or dietary patterns combined effects on ASCVD risk by ancestry

Figure 1 shows the effect of the high-risk PRS with each dietary pattern or high physical activity level on ASCVD risk. Among European Americans, high physical activity level was able to lower ASCVD risk by 13% in the second (RR=0.87;0.85-0.90) and highest (RR=0.87;0.84-0.90) PRS tertiles compared to the lowest PRS tertile. In addition, high physical activity level was able to lower ASCVD risk by 12%, when comparing those at the highest PRS tertile with high physical activity level vs. those at second PRS tertile who were physically inactive (RR= 0.88;0.85-0.90). Among African Americans, the influence of physical activity with the PRS showed 15% lower risk, seen only in the highest tertile compared to the lowest tertile (RR=0.85;0.80-0.90). Similarly for African Americans, physical activity had a 6% lower risk among those who were physically active in the highest PRS tertile compared to those in the second PRS tertile who were physically inactive (RR=0.94;0.89-0.98).

**Figure 1.**
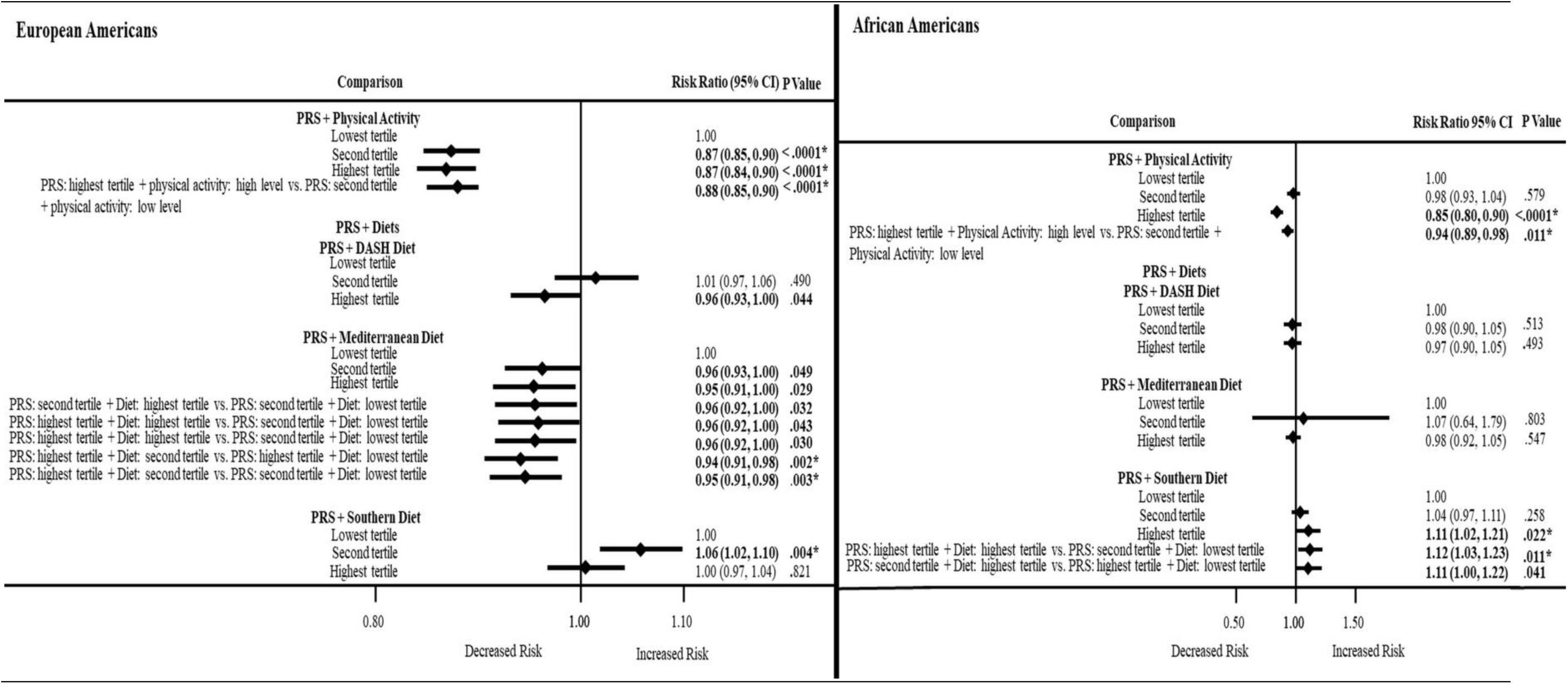
Association between a PRS, dietary patterns or physical activity. **Abbreviations:** PRS, polygenic risk score; DASH, Dietary Approached to Stop High Blood Pressure; **Bold indicates p values that were statistically significant at p < .05.** **\*Bonferroni adjustment for multiple testing (p < .05/2= < .025).** Atherosclerosis cardiovascular disease status was regressed against an interaction term composed of a principal component adjusted PRS, a dietary pattern score, and physical activity adjusting for a covariate propensity score composed of age, sex, current cigarette smoking status, current drinking status. Dietary pattern Z scores: Atherosclerosis cardiovascular disease status was regressed against dietary pattern Z scores (DASH, Mediterranean, and Southern diets) adjusting for a covariate propensity score composed of age, sex, physical activity, current cigarette smoking status, and current drinking status. Dietary pattern scores were computed using KMeans cluster, then converted to Z scores.

Figure 1 also shows ASCVD risks for the influence of the genetically high-risk PRS with each dietary pattern. Both the high diet quality DASH and Mediterranean dietary patterns are effective in European Americans at about 5% (RR=0.95;0.91-0.98) to 6% (RR=0.94;0.91-0.98), respectively in lowering ASCVD risk. However, these effects were not significant in African Americans. Among European Americans with a high PRS burden, the Mediterranean diet showed more ability to lower ASCVD risks than the DASH diet. However, only a few ASCVD risks (though significant), met Bonferroni adjustment due to multiple testing (p <.025). Among African Americans with a high PRS burden, the low-quality Southern diet had a profound effect with 11% (RR=1.11;1.02-1.21) to 12% (RR=1.12;1.03-1.23) higher ASCVD risk in the highest tertile compared to the lowest tertile, while European Americans had 6% (RR=1.06;1.02-1.10) higher ASCVD risk in second tertile compared to the lowest tertile.

### PRS, dietary patterns and physical activity combined effects on ASCVD risk by ancestry

Lastly, we examined the influence of the PRS with each the dietary pattern and high physical activity on ASCVD risk in participants with a high PRS burden (Figure 2). European Americans with a high PRS burden, had 11% to 15% lower ASCVD risks when they consumed a DASH or Mediterranean diet and had high physical activity. Likewise, in European Americans, high physical activity was able to lower the effects of the risk-raising PRS and the low-quality Southern diet by 9% (RR=0.91;0.85-0.96) to 15% (RR=0.85;0.80-0.90). However, African Americans had 13% to 18% lower ASCVD risks in the highest tertile compared to the lowest tertile for healthy DASH (RR=0.87; 0.78-0.97) and Mediterranean (RR=0.82;0.72-0.95) diets, respectively, attesting to the lowering effects of high physical activity on ASCVD risks in African Americans. In addition, in African Americans, there was a 20% lower ASCVD risk for the Southern diet (RR=0.80;0.64 to 0.99), but this effect did not meet Bonferroni correction for multiple testing.

**Figure 2.**
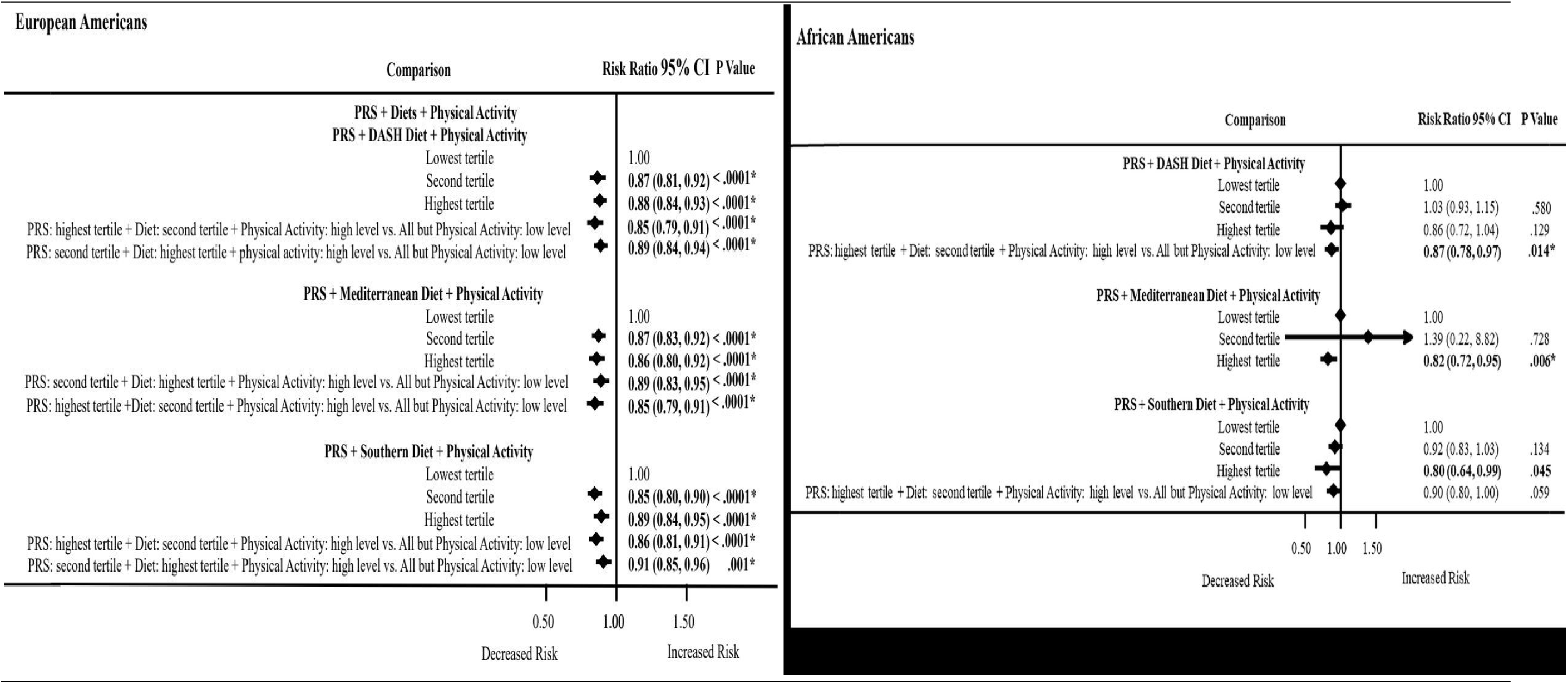
Association between PRS and physical activity or dietary patterns. **Abbreviations:** PRS, polygenic risk score; DASH, Dietary Approached to Stop High Blood Pressure. **Bold indicates p values that were statistically significant at p < .05.** **\*Bonferroni adjustment for multiple testing (p < .05/2= < .025).** PRS: Cardiovascular status was regressed against an interaction of the principal components adjusted PRS and physical activity or each dietary score adjusting for a covariate propensity score composed of age, sex, physical activity, current cigarette smoking status, and current drinking status. Te model with PRS by physical activity interaction did not include physical activity in the covariate propensity score. Dietary pattern scores were computed using KMeans cluster, then converted to Z scores.

### Interactions of the PRS, dietary patterns and physical activity on ASCVD risk by ancestry

Figures 3 and 4 show the predicted probabilities of ASCVD risk when a specific dietary pattern is consumed together with their level of physical activity across PRS tertiles from lowest to highest burden of risk-raising alleles. The relationships were clearer for European Americans than African Americans. The highest tertiles of DASH and Mediterranean diets with high physical activity, especially in European Americans, showed lower ASCVD risks. The Southern diet, as expected, raised ASCVD risk. However, those in the highest PRS burden were able to decrease their risk for ASCVD with a high physical activity. We did not find any mediation effects with dietary patterns on ASCVD risk.

**Figure 3.**
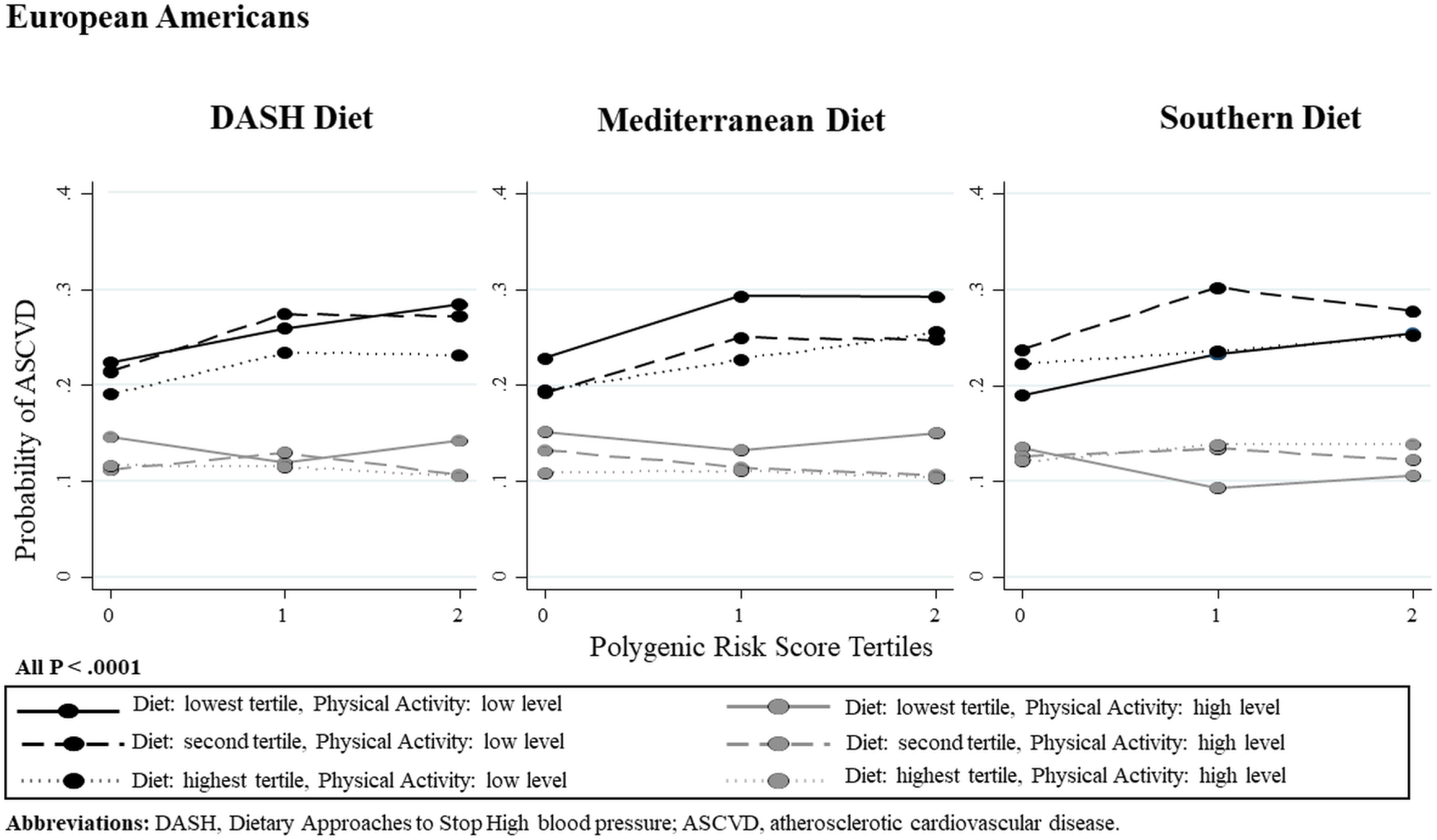
Predicted probabilities of ASCVD for consuming different levels of diets (DASH, Mediterranean, and Southern) within tertiles of a polygenic risk score. Summary p values for each line were calculated in metap in Stata using Fisher’s Exact test.

**Figure 4.**
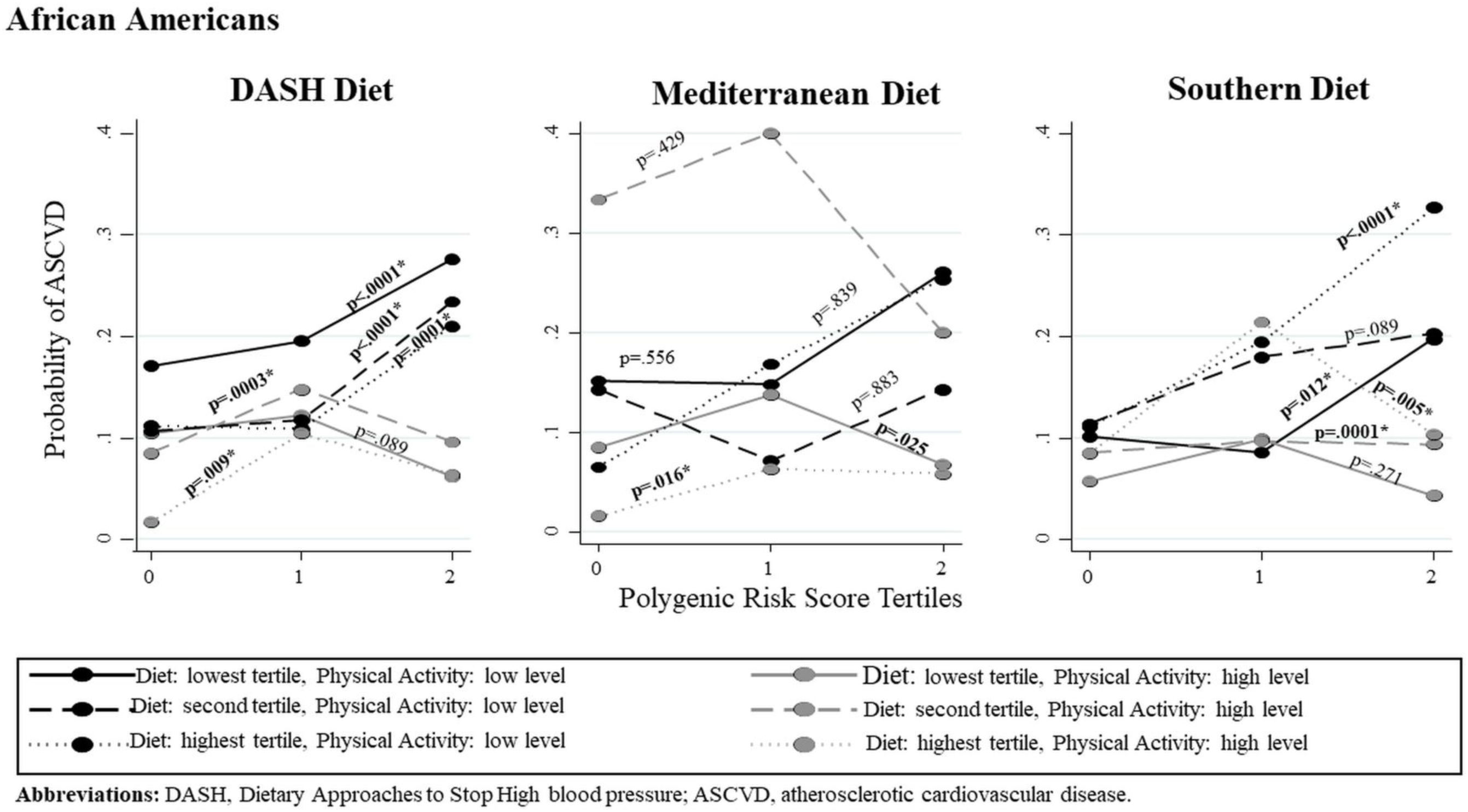
Predicted probabilities of ASCVD for consuming different levels of diets (DASH, Mediterranean, and Southern) within tertiles of a polygenic risk score. Summary p values for each line were calculated in metap in Stata using Fisher’s Exact test.

### Pathways of PRS mapped genes by ancestry

Among the 10 top pathways in European Americans and African Americans, genes for fructose metabolism and fructose catabolism were present (Figures 5 and 6). In addition, present were nicotinic acetylcholine receptors that allow both sodium and calcium ions and nutrient use and growth of ovarian cancer that involves glucose and glutamine utilization. In African Americans, pathways for Vitamin D strongly involved in depression; and dietary restriction and aging associated with cancer death.

**Figure 5.**
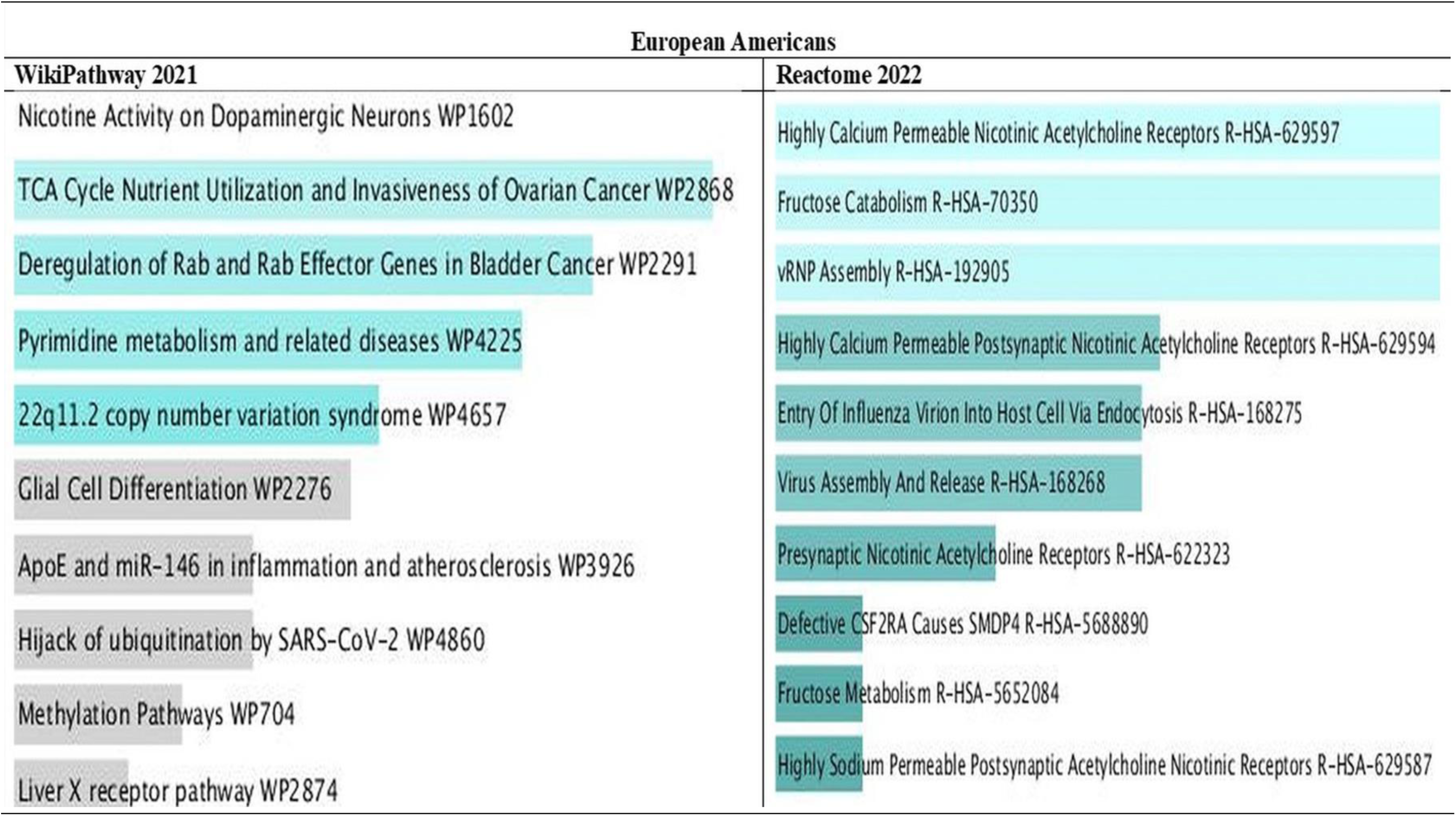
Bar chart of top pathways associated with atherosclerotic cardiovascular disease in European Americans for two libraries (WikiPathway and Reactome). The bar chart shows the top 10 enriched terms in the chosen library. Colored bars correspond to terms with significant combined p-values < 0.05 for Z score and the model p values. The brighter the color, the more significant that term is.

**Figure 6.**
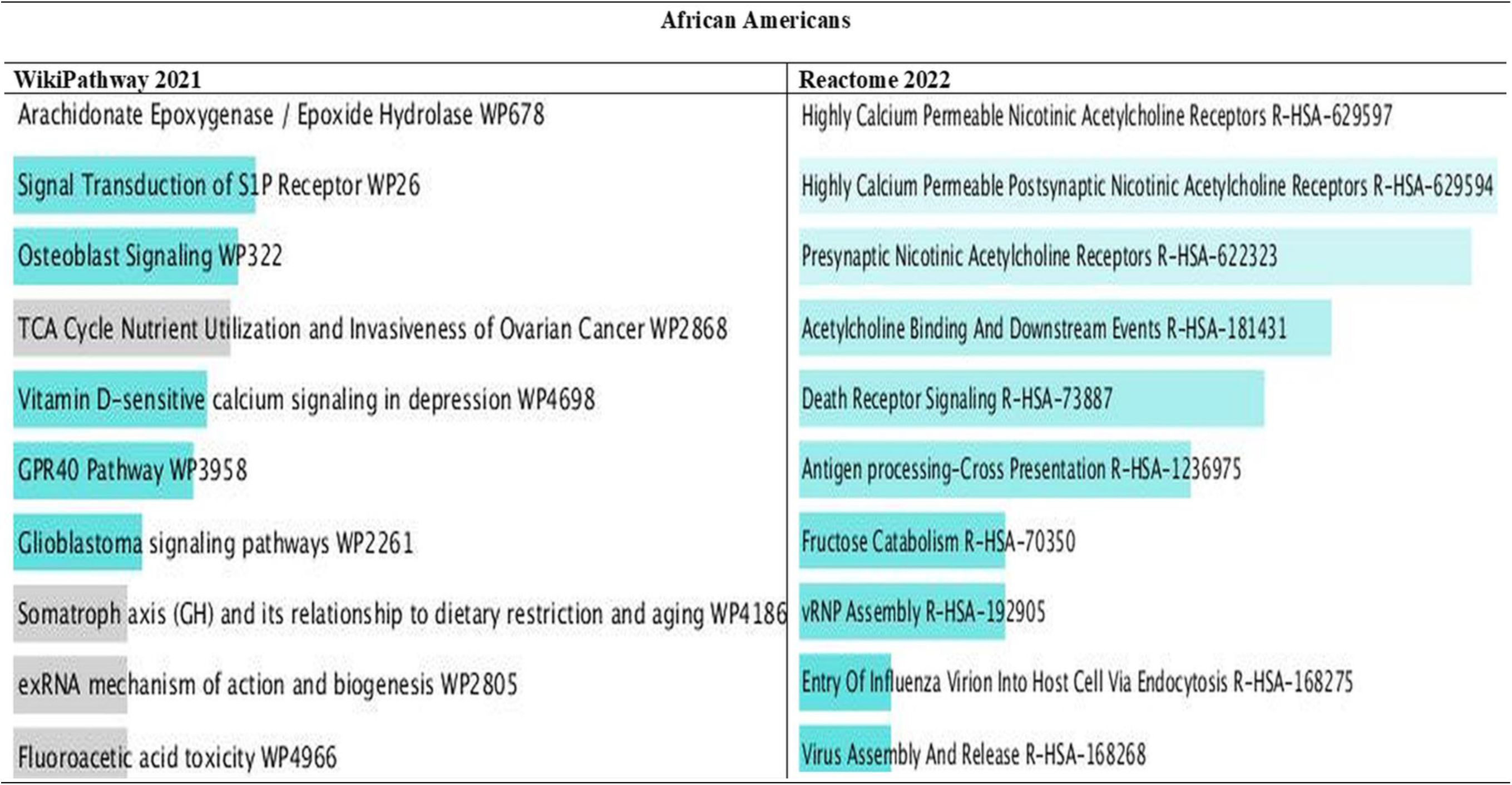
Bar chart of top pathways associated with atherosclerotic cardiovascular disease in African Americans for two libraries (WikiPathway and Reactome). The bar chart shows the top 10 enriched terms in the chosen library. Colored bars correspond to terms with significant combined p-values <0.05 for Z score and the model p values. The brighter the color, the more significant that term is.

## DISCUSSION

In this study, we found that a genetically high-risk PRS was associated with ASCVD in both European Americans and African Americans. African Americans tended to have higher magnitude of risks than European Americans but had more lowering effects from high physical activity. Among African Americans, the high-quality DASH and Mediterranean diets were not able to reduce the effects of the PRS by themselves; but among European Americans, there was 5% to 6% reduction in ASCVD risk. The low-quality Southern diet had 6% and 12% higher risk in European Americans and African Americans, respectively with a high PRS burden. High physical activity level alone was able to lower the risk-raising effects of the PRS by 13% in European Americans and by 15% in African Americans. However, lowering effects of high physical activity with the high-risk PRS and the high-quality DASH and Mediterranean diets and low-quality Southern diet lowered ASCVD risks by 9% to 15% in European Americans and in African Americans by 13% to 18% for the DASH and Mediterranean diets. There were differences in top pathways in both ancestries for fructose metabolism and catabolism and additional pathways for African Americans.

Awareness of one’s PRS has been shown to improve disease risks. Hasbani et al.^40^ reported that in their high-PRS burden group, an ideal Life Simple Seven score was associated with only 4.5 more CHD-free years compared with a poor Life Simple Seven score. Like Kurniansyah et al.^41^ who found that their hypertension-PRS was associated with coronary artery disease, ischemic stroke, type 2 diabetes, and kidney disease, our PRS was associated with ASCVD. In addition, our previous PRS was associated with type 2 diabetes.^42^ Similarly to Krapohl et al.^43^ who used the joint-effects of SNPs interaction in their PRS, we found that our joint-effects high-risk PRS was significantly associated with ASCVD risk.

Our results showed that the DASH and Mediterranean dietary patterns were able to reduce the effects of the PRS in European Americans and African Americans by a modest amount on ASCVD risk. In addition, when the participants with a high PRS burden engaged in habitual high physical activity, this together with a high-quality diet, showed a greater reduction in ASCVD risk. Furthermore, the effect of high physical activity was able to reduce the harmful effects of the risk-raising PRS-Southern diet combination by about 15% in European Americans and 20% in African Americans, but the latter did not meet Bonferroni correction possibly due to low sample size.

In the PRS-based pathway analysis, we found genes for fructose metabolism and fructose catabolism linked to high fructose corn syrup intake. Consumption of high fructose corn syrup promotes insulin resistance and obesity, elevated LDL cholesterol and triglycerides that can lead to metabolic syndrome.^44^ In addition, nicotinic acetylcholine receptors engage both sodium and calcium ions. High sodium intake is related to high blood pressure which is highly prevalent in African Americans. In African Americans, we found genetic-linked pathways for Vitamin D that are strongly related to depression. However, genetic-linked pathways for aging, abnormal metabolic effects, and metabolic syndrome can be reversed by intermittent fasting and dietary restriction.^45,46^ Molecular pathways that involve cancer genes associated with death signaling in African Americans should be further explored although some evidence showed that intermittent fasting may induce an anti-cancer response.^46^

PRSs are stable and inherent and can be used to estimate the risk of disease from an early stage or when the disease is not yet apparent. A genetically high-risk PRS may be beneficial in identifying high-risk individuals before a full formal risk assessment, and therefore can better allocate resources and reduce costs in primary care while still preventing ASCVD events.^47^ A PRS-diet-disease association can be modified by changes in diet and lifestyle factors. It can often become part of prevention programs and interventions. A genetically high-risk PRS can be a motivating factor to urge people to change their behaviors to decrease their chances of getting a disease. Because of the heritability of disease of everyone, interventions that consider patients’ genetic profile, risk factors and devise treatment plans to minimize further risk for disease development can be helpful in precision medicine.^48^

Our study has limitations. Our present study is a cross-sectional study in which only one time-point was used to access associations. In addition, because of the temporal nature of this study, we could not establish causal relationships, but could only establish associations between the PRS, dietary patterns and physical activity on ASCVD risk. An important observation to note is that the African American sample was much smaller sample than that of European Americans and, therefore, had less power to detect a statically significant association. Most of the significant statistics that passed Bonferroni correction (p < .025) were in the highest tertile compared to the lowest tertile. While European Americans frequently had more statistically significant estimates in the second and highest tertiles compared to the lowest tertiles. Nevertheless, we believe our results are an important contribution to science.

A major advantage of this study was that we were able to impute variants from the large 1000 genomes reference database to augment our sample of variants available for the PRS.^24^ In addition, we were able to combine data from seven rigorously performed NHLBI studies from dbGaP. Our sample observations decreased after dividing the genetic data into train and test samples. With the African Americans sample, though the sample size was small, we were able to discover important associations even when corrected for the Bonferroni adjustment. Recall bias and information bias were diminished because the protocols were established when the food frequency questionnaires and collection of other phenotypic information such as covariate information were administered together. In addition, we decreased bias by excluding outliers for total caloric intake in men and women. Furthermore, we decreased bias in food recall by dropping men and women who had outlier values that were above or below 1% percentile of total calories. Moreover, bias was diminished by converting the PRS and diets to Z-scores to standardize the dietary values across datasets.

Over the past several years, ultra-processed foods, an integral part of the Southern diet, have become a more common and acceptable source of nutrition, while intake of foods with higher diet quality have decreased. Studies have shown that ultra-processed foods increase the ASCVD risk.^49^ High quality diets such as DASH and Mediterranean diets can lower ASCVD risk, but only by a small to moderate proportion in European Americans with a genetically high-risk PRS. However, African Americans with a genetical high-risk PRS can benefit from a high physical activity level to decrease ASCVD risk. Individuals with a genetically high-risk PRS can therefore reduce their risk through appropriate lifestyle modifications. Because we found genetic-linked molecular pathways in African Americans related to Vitamin D and calcium that are strongly related to depression, and other pathways for aging and cancer death, reduction in ASCVD should be beneficial for African Americans to decrease the harmful effects of genes in top molecular pathways. Clinical trials and intervention studies are needed to investigate the responses of genetic effects and connected molecular pathways with diet quality. This study should be replicated in a larger sample of African Americans.

## Data Availability

The data is from dbGaP. It should be requested because it is not freely available.

https://www.ncbi.nlm.nih.gov/gap/

## Acknowledgments

The authors would like to thank Bamidele Tayo, PhD for his guidance in preparing datasets for genotyping imputation, and analysis of the imputed datasets for PRS preparation.

## Financial Disclosure

This work is supported by a K01 grant from the National Heart, Lung, and Blood Institute (NHLBI) grants: K01 HL127278 awarded to Dale Hardy, PhD. There was no additional external funding received for this study. The NHLBI had no role in study design, data collection and analysis, decision to publish, or preparation of the manuscript.

Datasets utilized in our study are from dbGaP. Their information is below.

| Datasets | Accession Numbers |
| --- | --- |
| ARIC | phg000035, phg000079, phs000280 |
| CARDIA | phg000091, phs000285 |
| CHS | phg000183, phs000287 |
| FHS OFFSPRING | phg000004, phs000007 |
| FHS GENX3 | phg000006, phs00007 |
| MESA | phg000071, phs000209 |
| WHI | phg000061, phs000200 |

---

**Supplemental Table 1.** Characteristics of diet forms used by the seven NHLBI Care studies.

| Study | Diet Form | Items | Exam Year Used |
| --- | --- | --- | --- |
| ARIC | FFQ | semi-quantitative 66-item | 1987-Exam 1 |
| CARDIA | Diet history | 102 questions | 1989-Exam 1 |
| CHS | Diet history | 96 questions | 1998-Exam 1 |
| FHS OFFSPRING | FFQ | semi-quantitative 88-items | 1983-Exam 3 |
| FHS GENX 3 | FFQ | semi-quantitative 88-items | 2002-Exam 2 |
| MESA | FFQ | semi-quantitative 153-items | 2010-Exam 5 |
| WHI | FFQ | semi-quantitative 145-items | 2005-Exam 1 |

**Abbreviations:** NHLBI, National Heart, Lung, and Blood Institute; Care, Candidate Gene Association Resource; ARIC, Atherosclerosis Risk in Communities study,<sup>29</sup> CARDIA, Coronary Artery Risk Development in Young Adults Study,<sup>33</sup> CHS, Cardiovascular Heart Study,<sup>34</sup> FHS, Framingham Heart Study Offspring and GENX 3 studies,<sup>31</sup> MESA, Multi-Ethnic Study of Atherosclerosis Study,<sup>30</sup> and Women's Health Initiative study (WHI).<sup>32</sup>

**Supplemental Figure 1.**
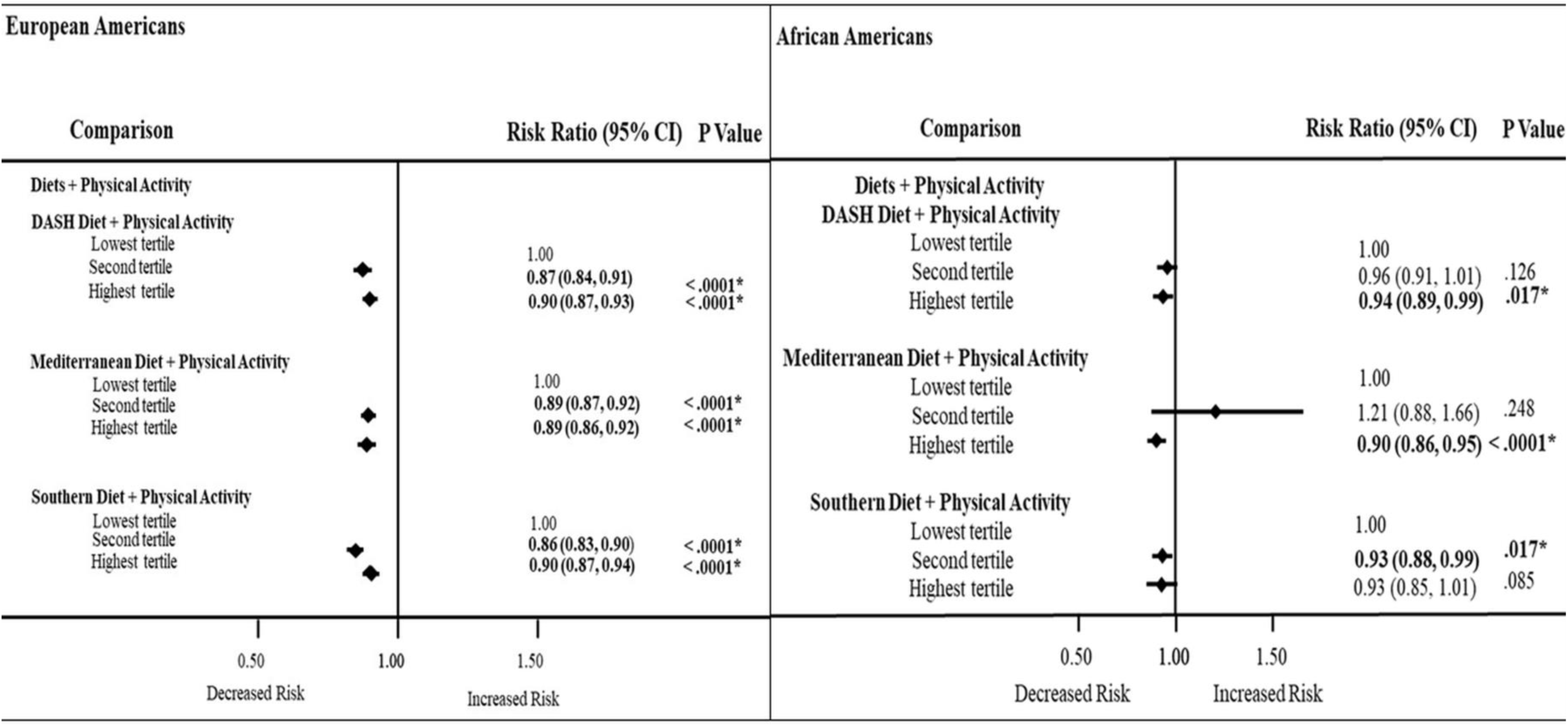
Association between dietary patterns and physical activity Abbreviations: DASH, Dietary Approached to Stop High Blood Pressure. **Bold indicates p values that were statistically significant at p < .05.** **\*Bonferroni adjustment for multiple testing (p < .05/2= < .025).** PRS: Atherosclerotic Atherosclerosis cardiovascular disease status was regressed against an interaction term composed of each dietary scores and physical activity adjusting for a covariate propensity score composed of age, sex, current cigarette smoking status, and current drinking status. Dietary pattern scores were computed using KMeans cluster, then converted to Z scores.

